# Top End Pulmonary Hypertension Study: Understanding Epidemiology, Therapeutic Gaps and Prognosis in Remote Australian Setting

**DOI:** 10.1101/2020.09.19.20197988

**Authors:** Pyi Naing, David Playford, Geoff Strange, Asanga Abeyaratne, Thomas Berhane, Sanjay Jospeph, Ellie Costelloe, Maddison Hall, Gregory M Scalia, Douglas L Forrester, Henrik Falhammar, Nadarajah Kangaharan

**Affiliations:** The Prince Charles Hospital; University of Notre Dame Australia; University of Notre Dame, Australia; Royal Darwin Hospital; Royal DarwinHospital; The Prince Charles Hospital; Karolinska Institute

**Keywords:** Pulmonary Hypertension, Indigenous, Remote Australia

## Abstract

**Introduction:** The Top End of Australia has a high proportion of Indigenous people with a high burden of chronic cardiac and pulmonary diseases likely to contribute to pulmonary hypertension (PH). The epidemiology of PH has not been previously studied in this region.

**Methods:** Patients with PH were identified from the Northern Territory echocardiography database from January 2010 to December 2015 and followed to the end of 2019 or death. PH was defined as a tricuspid regurgitation velocity ≥2.75 m/s measured by Doppler echocardiography. The etiology of PH, as categorized by published guidelines, was determined by reviewing electronic health records.

**Results:** 1764 patients were identified comprising 49% males and 45% Indigenous people. The prevalence of PH was 955 per 100,000 population (with corresponding prevalence of 1587 for Indigenous people). Hypertension, atrial fibrillation, diabetes and respiratory disease were present in 85%, 45%, 41% and 39%, respectively. Left heart disease was the leading cause for PH (58%), the majority suffering from valvular disease (predominantly rheumatic). Pulmonary arterial hypertension (PAH), respiratory disease related PH, chronic thromboembolic PH (CTEPH) and unclear multifactorial PH represented 4%, 16%, 2% and 3%, respectively. Underlying causes were not identifiable in 17% of the patients. Only 31% of potentially eligible patients were on PAH-specific therapy. At census, there was 40% mortality, with major predictors being age, ePASP and Indigenous ethnicity.

**Conclusion:** PH is prevalent in Northern Australia, with a high frequency of modifiable risk factors and other treatable conditions. Whether earlier diagnosis, interpretation and intervention improves outcomes merits further assessment.

## 1. Introduction

Pulmonary hypertension (PH) is a debilitating medical condition with a poor prognosis. It is not a single disease process, but rather a haemodynamic sequalae of several underlying pathologies. There are five distinct PH classification groups [1], and the management and prognosis of each is very different [2, 3]. PH is traditionally defined as mean pulmonary artery pressure (mPAP)≥25mmHg, measured at rest via right heart catheterization (RHC) [2]. This definition of PH has been changed recently with the pressure reduced to ≥20mmHg [1], due to data suggesting mildly elevated pulmonary artery pressures may be associated with increased mortality [4, 5]. In clinical practice, echocardiography provides the first objective evidence of PH and other valuable information such as presence or absence of left heart pathology. It may also provide evidence of increased pulmonary vascular resistance (PVR), a necessary parameter to diagnose pulmonary arterial hypertension (PAH), the only group of PH with evidence based specific PH therapy [6]. Despite the development of multiple advanced PH therapies over the last two decades [7], the mean survival of PH patients is approximately four years from the first recorded elevated pulmonary pressure by echocardiography [8, 9]. Emerging data suggest that the prevalence of all types of PH is rising [8, 10]. There was a recent report from Central Australia investigating its epidemiology [11]. We sought to evaluate PH in the Top End of Australia where nearly two-thirds of the Northern Territory (NT) population live and have access to diagnostics and therapy through a tertiary hospital.

### 2. Background and Study Rationale

The medical service delivery is challenging in the Top End of Australia due to *tyranny of distance* for many residents, limited health resources and health inequity faced by the Aboriginal and Torres Straits Islanders who represent approximately 27% of the population [12]. Non-communicable chronic diseases (e.g. coronary artery disease, hypertension, diabetes) as well as communicable chronic diseases (e.g. rheumatic heart disease) are highly prevalent in the region [13]. These may lead to PH and subsequent burden of morbidity and mortality in the absence of or delay in management of underlying pathology. Advanced PH therapy is recommended for the treatment of PAH, however it requires specialist centre prescription, careful monitoring and is expensive [14]. Moreover, there are significant delays in definitive diagnosis of PH leading to delayed commencement of appropriate therapy [15, 16]. Better understanding of PH epidemiology will allow better planning and management of scarce health resources and thence hopefully improve the outcomes. We have a unique advantage of studying epidemiology of PH in the Top End as a single cardiology service, NT Cardiac Pty Ltd, provides the echocardiography service to the whole region. This improves data capture and integrity not only from the point of view of individual cases but also the availability of all cardiac investigations for review.

We hypothesized that the prevalence of PH in the Top End is higher than elsewhere in Australia and potentially contributes to the life expectancy gap between Indigenous and non-Indigenous Australians. Hence, we conducted this cohort study to better document the epidemiology, associated comorbidities and prognosis of PH in this region.

## 3. Methods

### 3.1. Ethics, study design and data sources

Ethics clearance was obtained from the Human Research Ethics Committee of the NT Department of Health and Menzies School of Health Research. Structured Query Language (SQL) queries were made in the NT Cardiac echo-database to identify echocardiograms with maximal tricuspid regurgitation velocity (TRV_max_)≥2.75m/s, which is equivalent to estimated pulmonary artery systolic pressure (ePASP)≥40mmHg assuming a right atrial pressure of 10mmHg, a methodology consistent with previous studies [8, 17]. Only residents of the Top End were included. Data were collected from echocardiograms performed between January 2010 and December 2015 and the patients were followed up until death or the end of 2019, whichever occurred first. When multiple echocardiograms were found for the same patient, the first echocardiogram was used to determine the classification and severity of PH. Corresponding clinical information were collected from electronic medical records (EMRs) from both the Department of Health and NT Cardiac. The date of diagnosis was defined as the first date an echocardiogram showed evidence of PH which could be the date before January 2010. Survival and follow-up time were calculated as the time difference between the date of diagnosis and death or census date. Patient’s place of residence was determined by the current post code. The Strengthening the Reporting of Observational Studies in Epidemiology (STROBE) guidelines in reporting were strictly followed [18].

### 3.2. Study Setting

Top End is the upper half of the NT and contains four main regions: Darwin, Kakadu national park, Arnhem land and Katherine region. It covers an area of approximately 245,000 km^2^ and had a population of 184,657 according to 2016 census data. The cardiology service is provided by NT Cardiac based in Darwin with regular outreach specialists’ visits to remote communities and to the two regional hospitals (Katherine and Gove District Hospital).

### 3.3. Determining the PH classification

A diagnostic algorithm slightly modified from the current guidelines was used in classifying patients into PH groups as illustrated in figure 1 [2]. The indexed echocardiogram report was first reviewed to look for left heart pathologies defined as moderate to severe left sided valvular (aortic and mitral) disease and left ventricular (LV) systolic or diastolic dysfunction (figure 1). These were determined by the reporting cardiologist in accordance with the American Society of Echocardiography (ASE) guidelines [19-21]. The presence of congenital heart disease was also identified in the echocardiogram report. Presence of significant (moderate to severe) respiratory pathologies (sleep apnoea, obstructive and restrictive lung disease) were determined by extensive review of the EMRs. Further causes of PH such as portal hypertension, chronic kidney disease and connective tissue diseases were also identified in the EMRs. If more than one cause of PH was present for one patient, the most severe pathology was recorded as the etiology of PH. Patients were classified as PAH only if RHC was performed and pre-capillary PH physiology (Pulmonary artery wedge pressure (PAWP)≤15mmHg and PVR>3WU) were demonstrated, and no alternative causes were identified. A few exceptions to this rule were patients who had evidence of portal hypertension who could be classified as portopulmonary hypertension (PPHTN), and patients with congenital heart diseases known to cause PAH. Patients without any identifiable pathology were labelled as PH with unidentifiable cause, only after exhaustive review of all available records.

**Figure 1.**
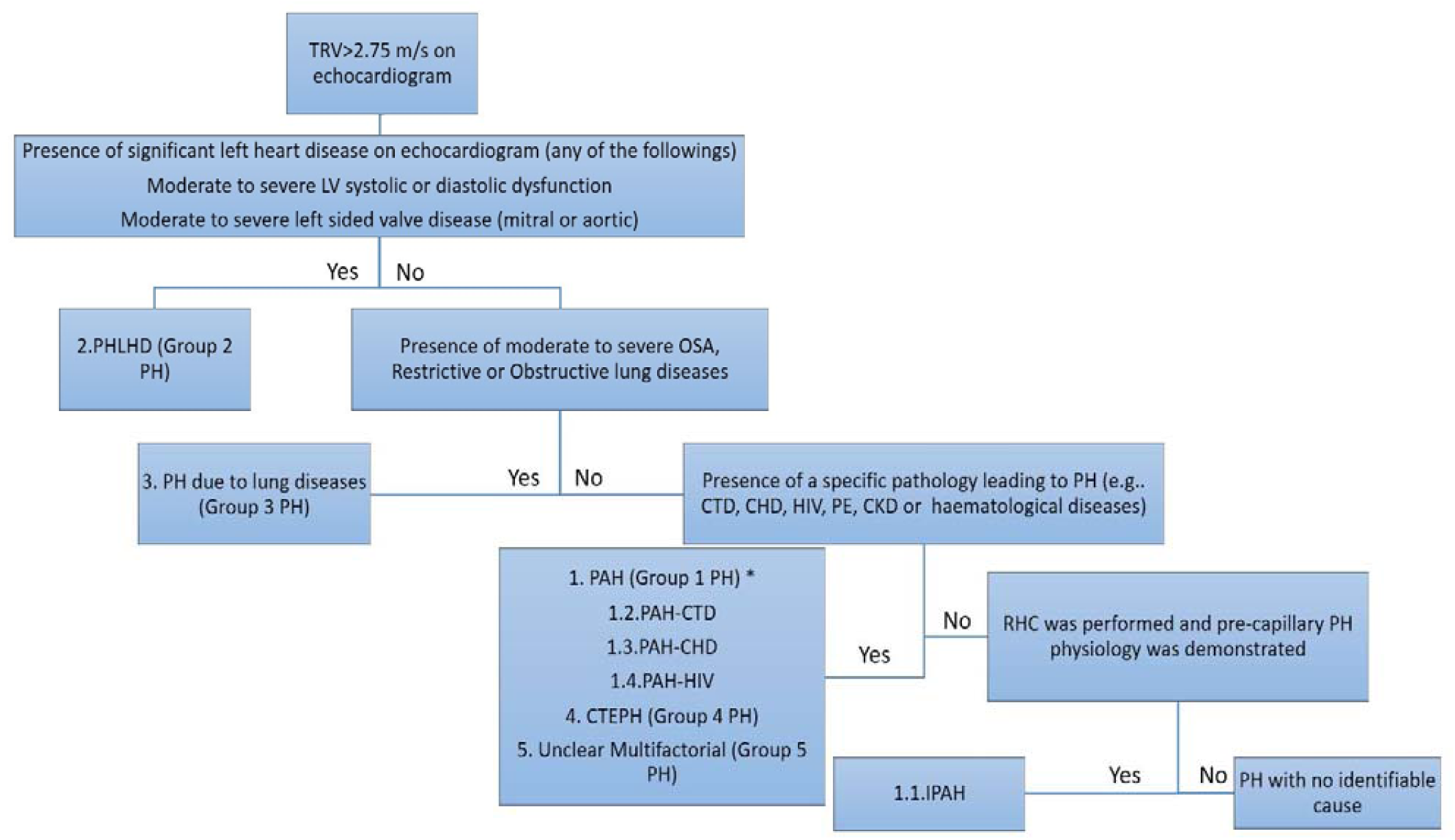
Diagnostic algorithm used in the study. TRV= tricuspid regurgitation velocity, LV= left ventricle, PH-LHD= pulmonary hypertension due to left heart disease, PH= pulmonary hypertension, OSA= obstructive sleep apnoea, PE= pulmonary embolism, HIV= human immunodeficiency virus infection, CTD= connective tissue disease, CKD= chronic kidney disease, CHD= congenital heart disease, PAH= pulmonary artery hypertension, CTEPH= chronic thromboembolic pulmonary hypertension, IPAH= idiopathic PAH, RHC= right heart catheterisation, *if RHC was performed and pre-capillary PH physiology was demonstrated except in patients with portal hypertension and CHD that can cause PAH

### 3.4. Statistical Methods

Statistical analysis was performed using International Business Machines Statistical Package for the Social Sciences (IBM® SPSS) 26.0 software. The continuous variables were reported as mean (± standard deviation) or median (interquartile range/IQR) as appropriate. Pearson’s chi-square test was used to compare the categorical variables while Student’s t-test was used to compare the means. Man-Whitney U test was used to compare the medians. The Kaplan-Meier method was used for survival analysis. Cox regression analysis was performed to determine factors associated with increased mortality.

## 4. Results

### 4.1. Study Cohort and overall prevalence of pulmonary hypertension

There were total of 46,731 echocardiograms performed during the study period. Initially 2072 echocardiograms were identified after removing duplicates with the same hospital record numbers and those performed in Central Australia (Figure 2). After resolution of duplicates and exclusion of patients who were not residents of Top End and two patients with missing clinical data, the final cohort for analysis consisted of 1764 patients. Thus, the prevalence of PH in the Top End was 955 per 100,000. The prevalence for the Indigenous population was 1587 per 100,000 population (791 PH patients for 49,857 Indigenous inhabitants calculated as 27% of 184,657 from 2016 census).

**Figure 2.**
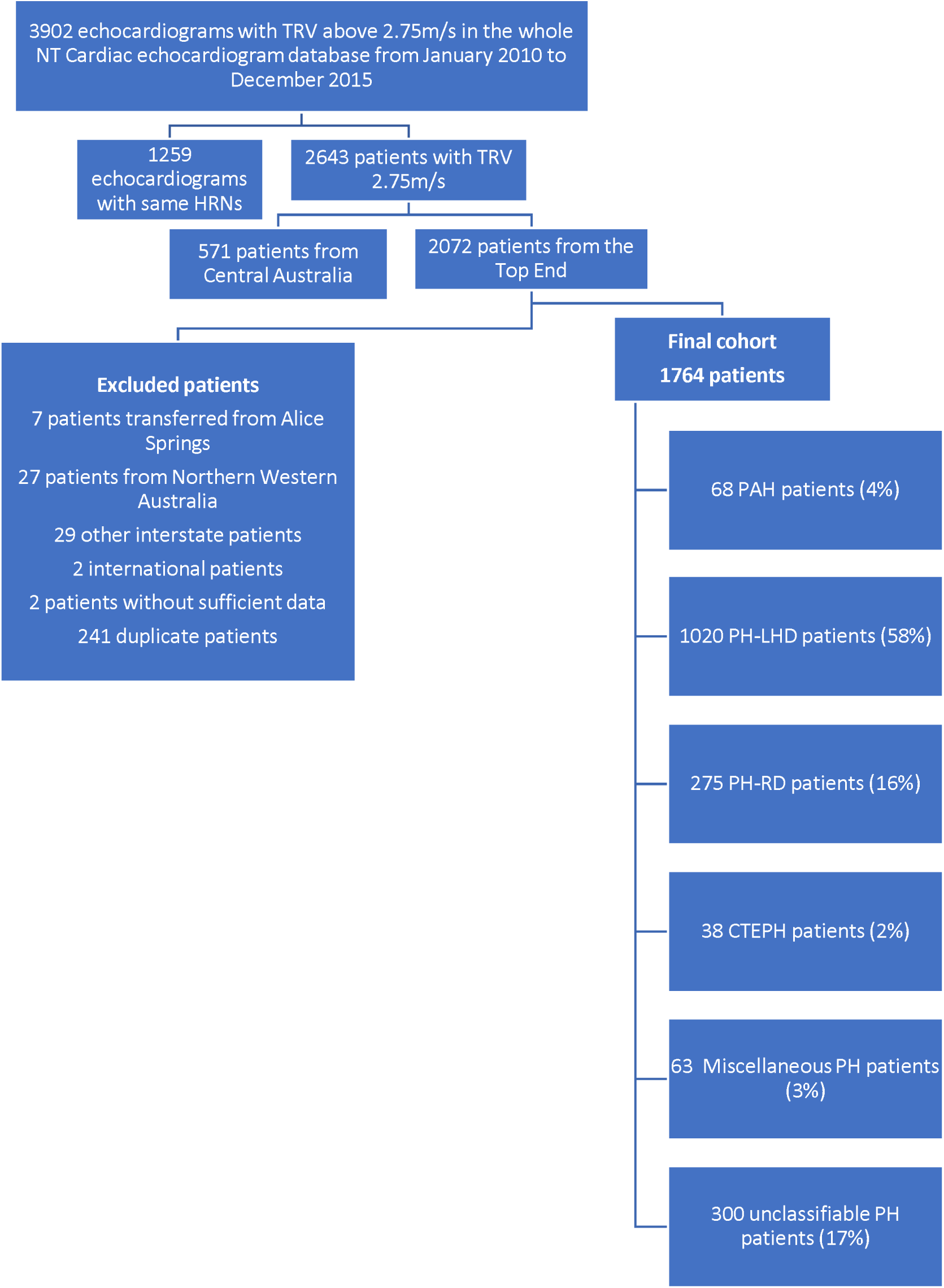
The study flow chart. HRN = Hospital Record Number. PAH = Pulmonary Arterial Hypertension, PH-LHD = Pulmonary Hypertension due to left heart disease, PH-RD = Pulmonary Hypertension due to respiratory disease, CTEPH = Chronic Thromboembolic Pulmonary Hypertension.

### 4.2. Clinical characteristics and comorbidities

The study cohort was evenly distributed between biological sex (49% male). Indigenous patients represented 45% (N=791) of the study population while making up 27% of the total population. The mean age of the entire cohort was 60±18 years. Indigenous patients were younger at diagnosis with a mean age of 48±18 years compared to 69±14 years for non-Indigenous patients (P=<0.001). This trend was true across all clinical classification groups as shown in Figure 3. The cohort contained 62% mild (PASP 40-49 mmHg), 22% moderate (PASP 50-59 mmHg) and 16% severe PH (PASP ≥60 mmHg) patients. The chronic diseases and modifiable risk factors were highly prevalent as illustrated in Table 1. Among the risk factors, smoking, diabetes, chronic kidney disease and left heart disease were more prevalent in Indigenous patients while atrial fibrillation and obstructive sleep apnoea were more common in non-Indigenous patients. There were 33 patients younger than 18 years of age in the cohort. The majority (n=26) was due to left heart disease and rheumatic heart disease was the underlying cause in 21 out of 26 patients. Six young patients had congenital heart disease. The cause was not identifiable in one child with PH.

**Table 1.**
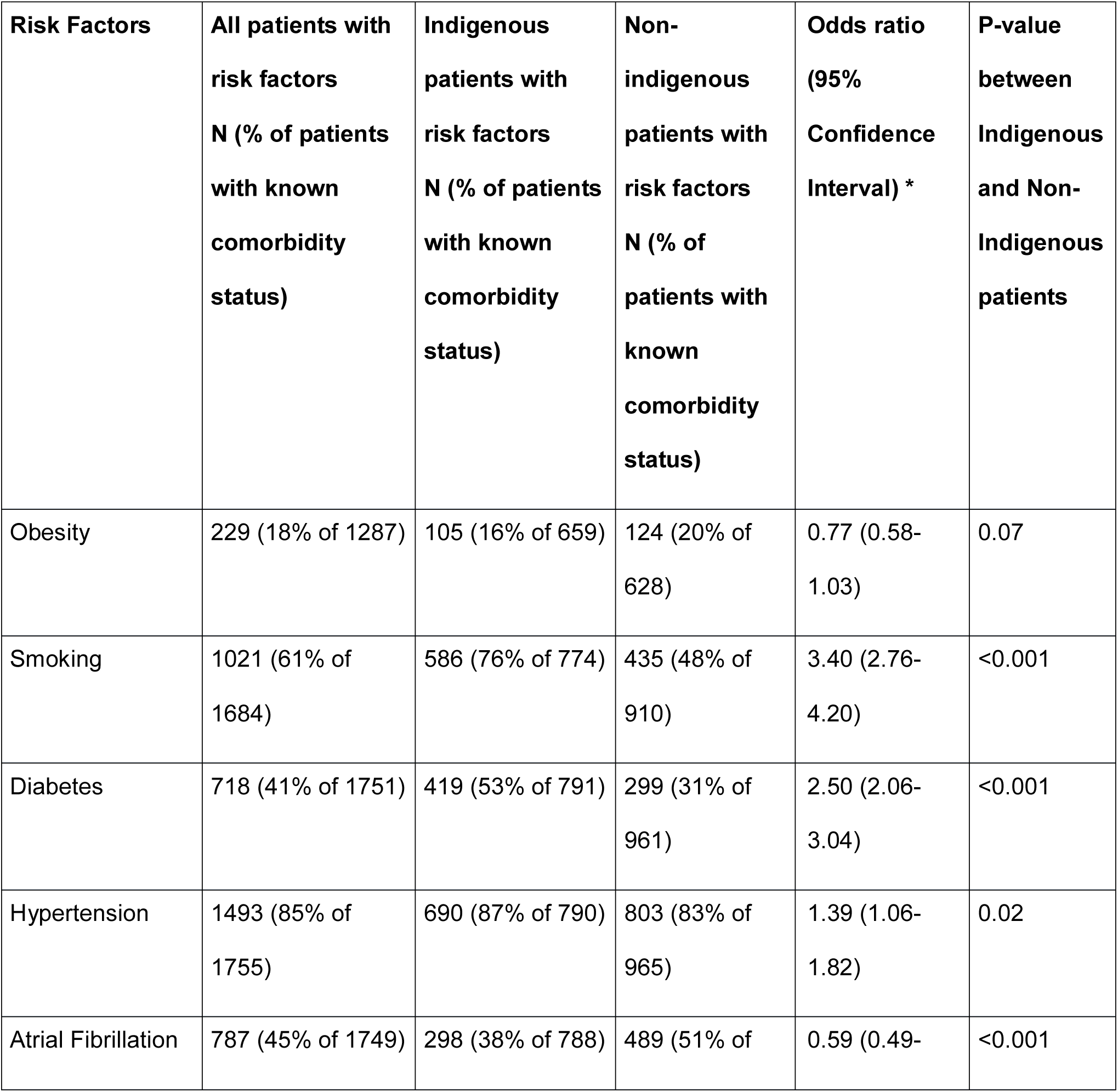
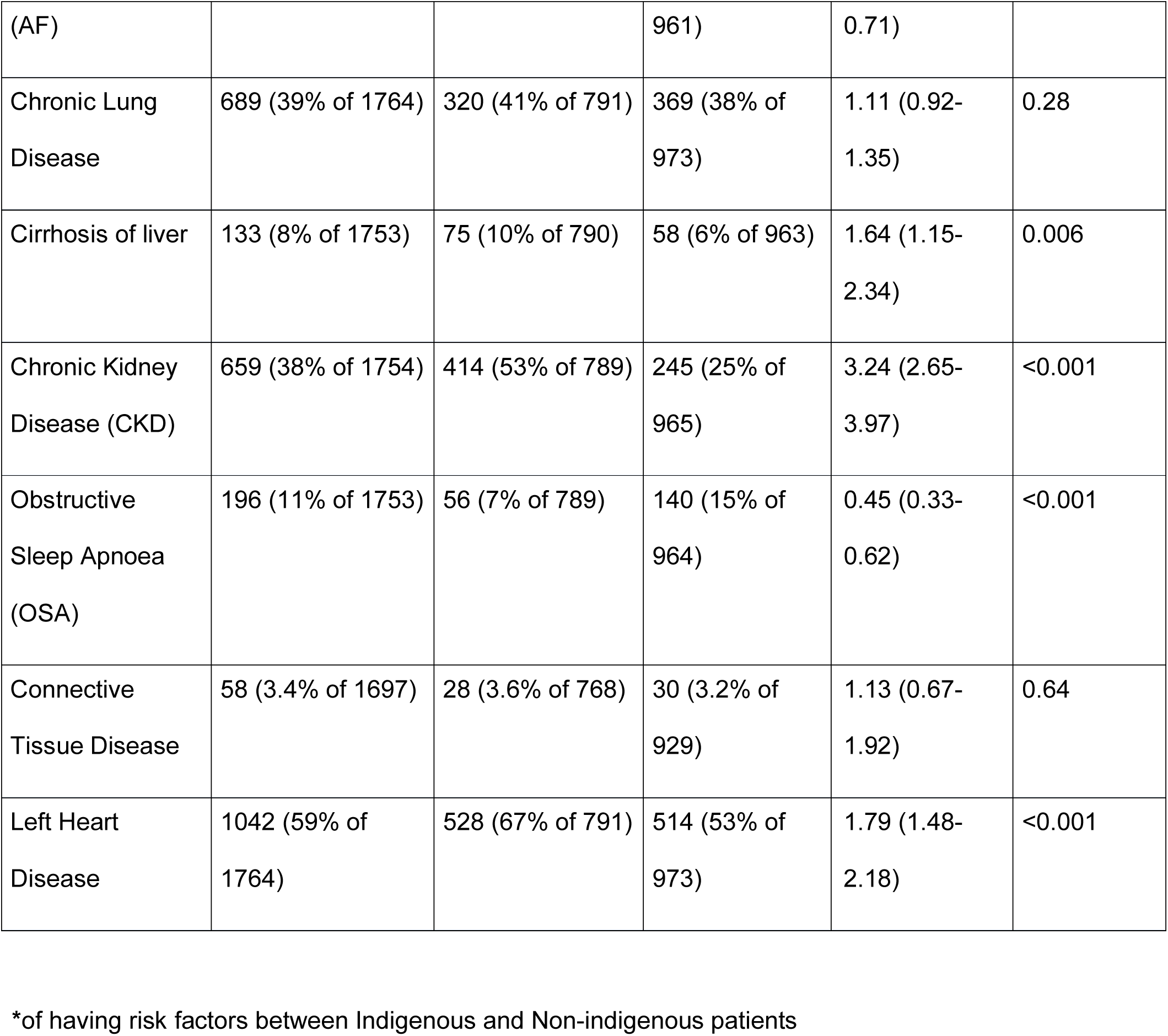
The frequency of risk factors and chronic diseases in the study cohort of patients living in the Top End of Australia with pulmonary hypertension

**Figure 3.**
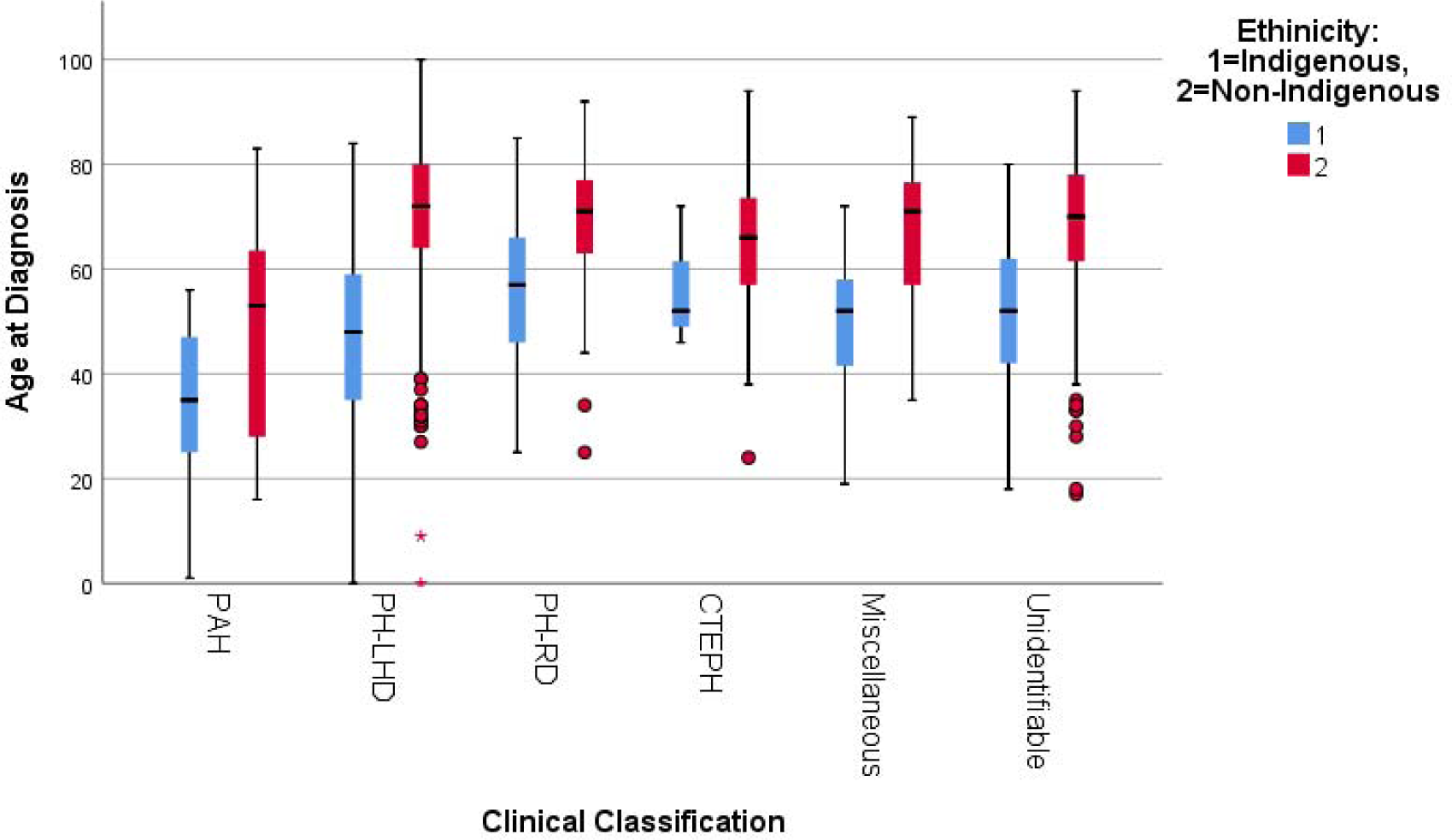
The box and whisker chart illustrating median age at diagnosis (the solid bar), interquartile range (the box) and highest and lowest values (the whiskers) and the outliers (the circles) according to different clinical classification of pulmonary hypertension (PH). Indigenous patients were younger at diagnosis in all PH classification groups. PAH= Pulmonary Arterial Hypertension, PH-LHD= Pulmonary Hypertension due to left heart disease, PH-RD= Pulmonary Hypertension due to respiratory disease, CTEPH= Chronic Thromboembolic Pulmonary Hypertension. P-value= <0.001 when comparing the median age at diagnosis for all Indigenous patients [49 (37-60)] years with Non-indigenous patients [71 (62-78 years)] using Mann-Whitney U test.

### 4.2. Differences among the PH groups

The comparison of demographics among the different PH groups is illustrated in Table 2. PAH was responsible for 4% of the cohort. PH due to left heart disease (PH-LHD) was the predominant group, making up 58% of the total cohort. Respiratory disease related PH was accounted for 16% of the patients while chronic thromboembolic PH (CTEPH) and unclear, multifactorial PH were responsible for 2% and 3% of the patients, respectively. The etiology of PH was not identifiable in 17% of the patients despite thorough search of the available medical records.

**Table 2.**
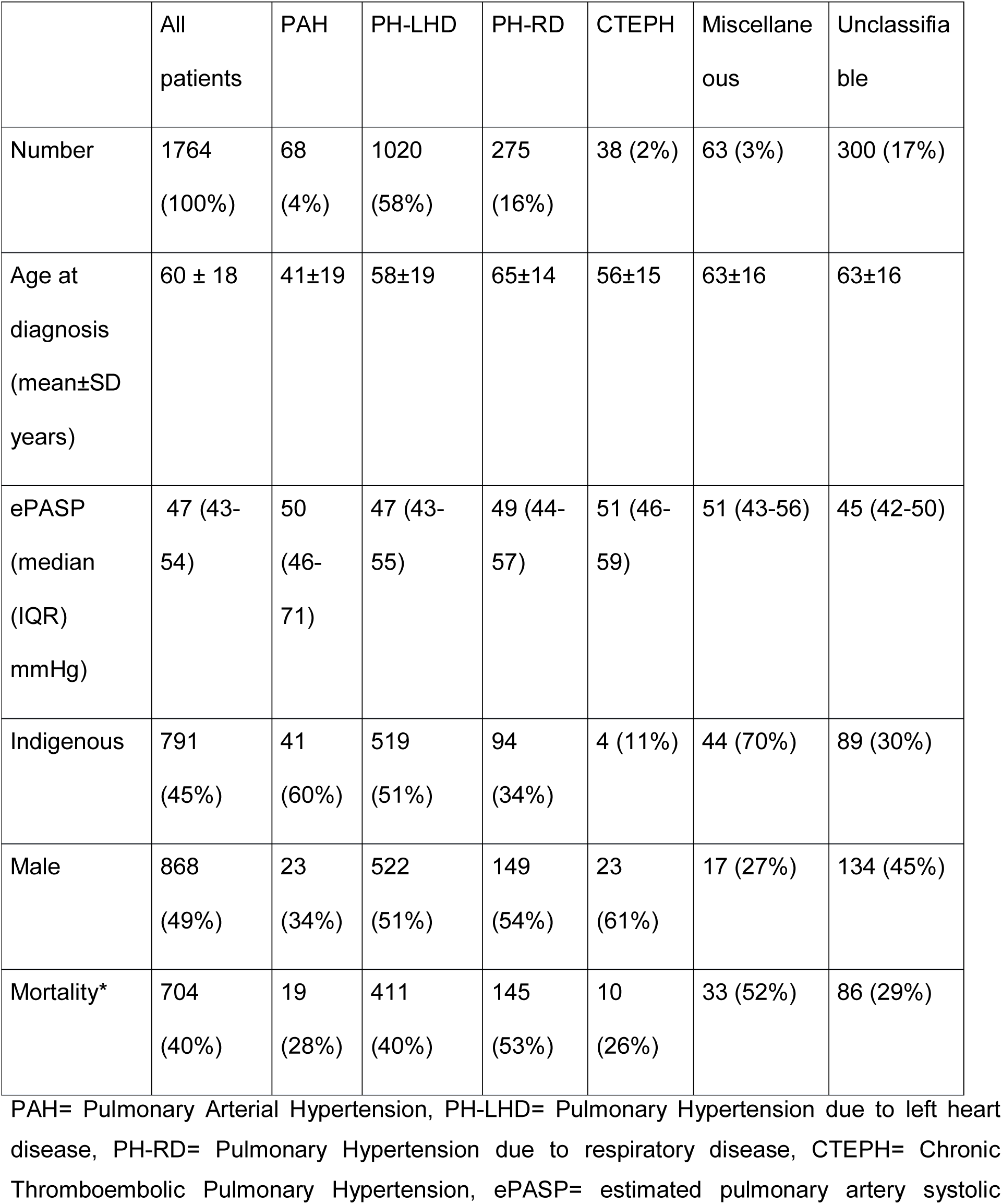
Demographics of patients in the study cohort, living in the Top End of Australia, according to the clinical classification of Pulmonary Hypertension

Valvular heart disease was responsible for 55% of patients with PH-LHD while left ventricular systolic dysfunction and diastolic dysfunction were responsible for 29% and 16% of PH-LHD patients, respectively. Left atrial size was reported to be dilated in 849 out of 982 PH-LHD patients (86%) where left atrial size was documented. Among PH patients with unidentifiable cause, 48% were reported to have dilated left atria. PAH patients were generally younger at diagnosis with a mean age of 41±19 years.

### 4.4. PAH patients and advanced PH therapy

There were four iPAH patients among the PAH group. Congenital heart disease was the most common cause of PAH in the cohort with 43%, followed by connective tissue disease related PAH (PAH-CTD) with 29%. Systemic lupus erythematosus (SLE) related PH was almost exclusive to Indigenous women (13 out of 16 patients) while no Indigenous patient was diagnosed with systemic sclerosis. All four iPAH patients received dual therapy with an endothelin receptor antagonist (ERA) and a phosphodiesterase 5 (PDE5) inhibitor. PPHTN due to cirrhosis of liver was the underlying etiology for 15 patients (22% of PAH patients). Only one PPHTN patient received advanced PAH-specific therapy. Overall, only 21 out of 68 patients (31%) with PAH received one form of advanced therapy.

### 4.5. Mortality data and predictors of increased risk

At the end of 2019, with a median (IQR) follow-up of 5.1 (2.9-7.4) years, there were 40% of patients with confirmed mortality (Figure 4). PH-RD was associated with the highest mortality, closely followed by PH caused by unclear, multifactorial causes. PH-LHD patients also suffered a high mortality with approximately 30% mortality at 5 years from diagnosis. PAH patients experienced the best prognosis in our cohort despite the low percentage of them receiving advanced therapy. Cox regression survival model analysis revealed that age at diagnosis (0.03% increase in risk for every year increase in age) and ePASP (0.012% increase in risk for every 1mmHg increase) were significant predictors of mortality. There was no gender-based difference in mortality. Indigenous status was associated with increased mortality (HR=1.796 (1.496-2.155), P=0.001) (Table 3 and Figure 5).

**Table 3.** Different variables predicting increased mortality in patients with pulmonary hypertension living in the Top End of Australia

|  | Hazard Ratio (95% CI) | P-values |
| --- | --- | --- |
| Age at Diagnosis | 1.033 (1.027-1.039) | 0.001 |
| Male gender | 1.123 (0.967-1.306) | 0.129 |
| Indigenous status | 1.796 (1.496-2.155) | 0.001 |
| ePASP | 1.012 (1.006-1.018) | 0.001 |

CI= confidence interval.

**Figure 4.**
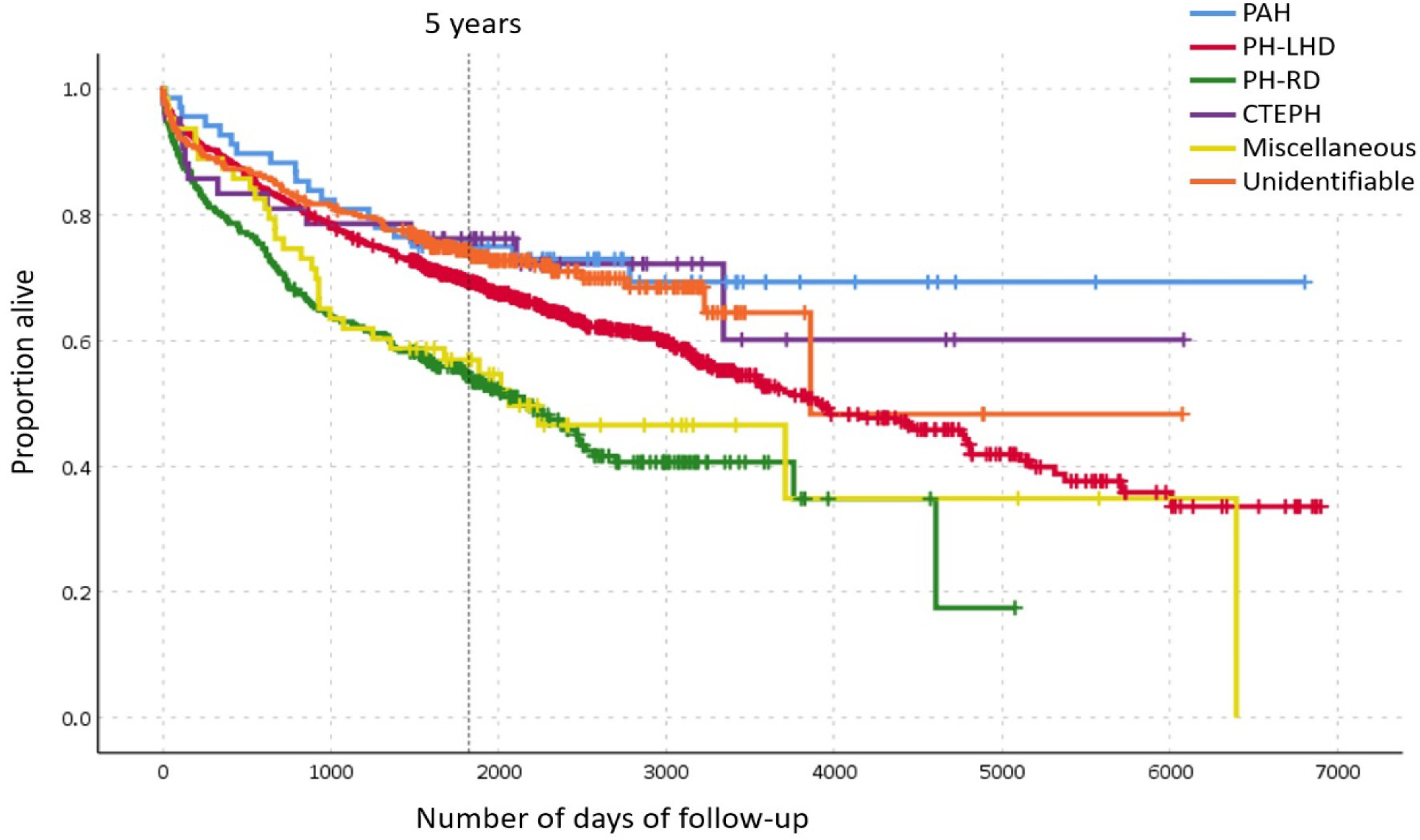
Kaplan-Meier Survival curves depicting the survival of patients according to the clinical classification of PH. PH-RD was associated with the worst prognosis. PAH= Pulmonary Arterial Hypertension, PH-LHD= Pulmonary Hypertension due to left heart disease, PH-RD= Pulmonary Hypertension due to respiratory disease, CTEPH= Chronic Thromboembolic Pulmonary Hypertension

**Figure 5.**
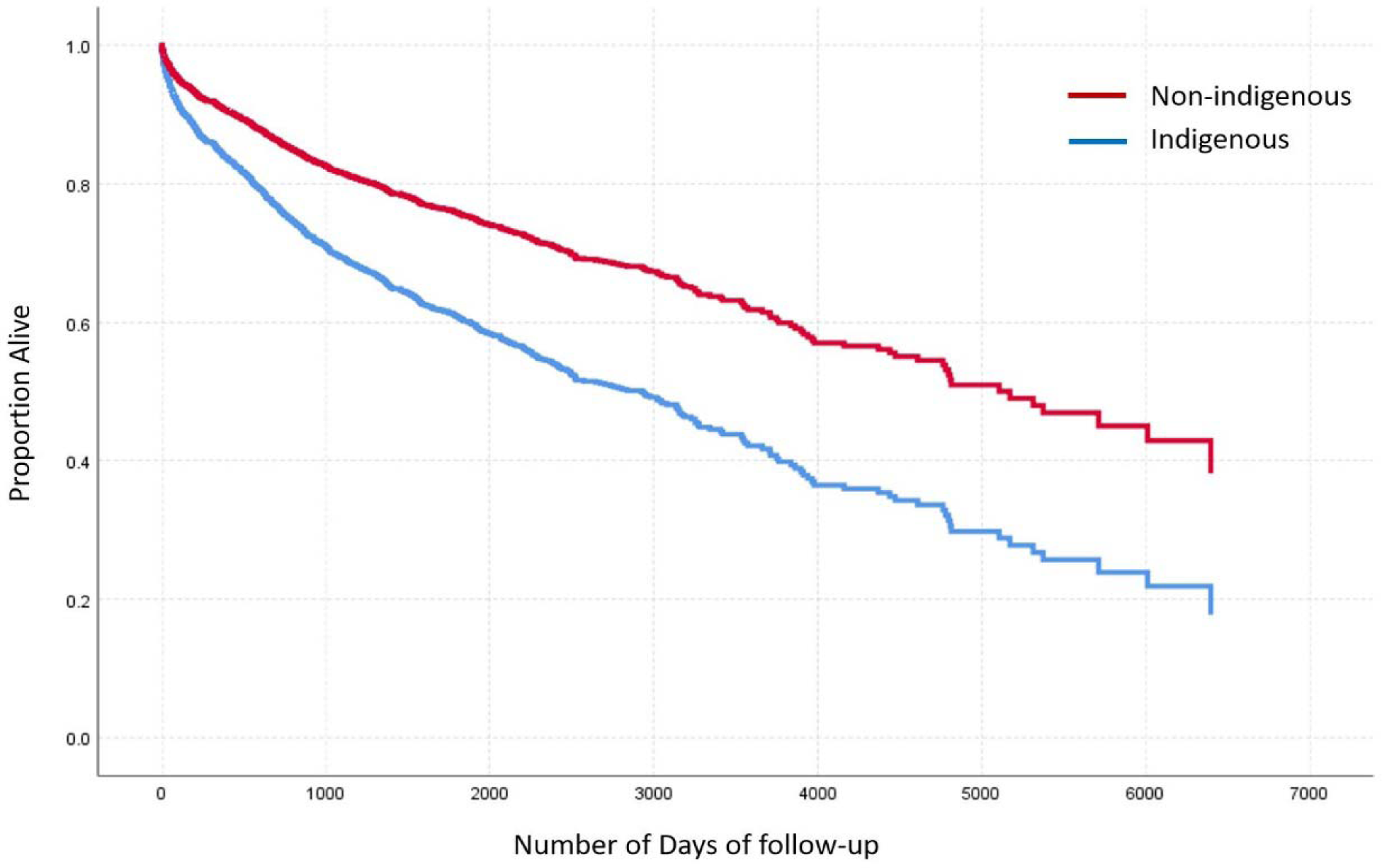
The survival curves of Indigenous and non-Indigenous patients constructed using multivariate Cox regression analysis. Hazard ratio=1.796 (1.496-2.155), P=<0.001. Covariates = age at diagnosis, gender and estimated pulmonary artery systolic pressure (ePASP).

## 5. Discussion

This is the most comprehensive description of PH in the Top End of Australia to date. Our study provides rich and comprehensive information regarding the prevalence and the root causes of PH in our region and complements the findings of the previously published Central Australian Study to complete the picture for the whole NT. PH was common, with a prevalence of approximately 1% of the total population in the Top End and was associated with a high mortality. The Indigenous population not only had a higher prevalence (1.6%), but also a higher mortality risk associated with PH. They were also diagnosed with PH at younger age.

The major strengths of our study were the integrity of the dataset due to the coverage of the area by a single echocardiographic database and all echocardiograms performed by certified, experienced cardiac sonographers and reported by experienced cardiologists. A limitation of our study was that we described the minimum predicted prevalence as the data was retrospectively obtained from an echocardiography database rather than prospectively screening the whole population. We may have also missed PH patients with insufficient tricuspid regurgitation to estimate PASP due to our study design, a finding highlighted by other studies of this type [8]. The RHCs were performed only in 92 patients (5.2% of the study cohort). This limitation may be the reason that the underlying cause was not found in a large portion (17%) of our cohort. Without confirmation of elevated PVR by RHC, the diagnosis of precapillary PH or PAH cannot be made and patients would not be able to access effective advanced PH therapies [1, 2]. Nonetheless, elevated pulmonary pressure estimated from TRV_max_ in echocardiography show an important signal towards a higher prevalence of PH. On the other hand, using TRV_max_ alone without other echocardiographic signs suggestive of PH in patients with intermediate probability of PH (TRVmax 2.9 to 3.4 m/s) may overestimate the prevalence in our study [2]. Many diagnoses and classification for the studied patients were “presumptive” and based on retrospective data. However, they highlight the important nature of this set of diseases and the need for comprehensive evaluation of every patient with pulmonary hypertension suspected on echocardiography.

There were different risk factor profiles between the Indigenous and non-Indigenous patients and specific tailored interventions may be considered to reduce these risk factors in both populations. The burden of risk factors and comorbidities were high in this region where approximately 30% of population was Indigenous and had high prevalence of rheumatic heart disease. We found that left heart disease was the leading cause of PH in our study in agreement with previous reports [8, 22]. Most patients in this group had valvular heart disease which likely depicts the ongoing public health problem of high prevalence of rheumatic heart disease in our region. Although PAH-CTD patients represented only small proportion of the cohort, there were specific demographic groups within this cohort, e.g., SLE-PAH being almost an exclusive disease of Indigenous women and no systemic sclerosis being diagnosed in Indigenous population. The remote, rural, Indigenous population also continue to suffer from preventable childhood PH caused by rheumatic heart disease in high numbers.

The prevalence of all types of PH in Top End was approximately three times higher than observed in the Armadale echo study [8] and higher than reported from Central Australia [11]. Epidemiology of PH worldwide is highly heterogenous creating challenges to clinicians and health authorities trying to tackle this emerging health problem. A function of reducing the pressure criteria for the haemodynamic definition of PH will result in a significant a priori increase in the prevalence of PH [1]. The hope is that this may lead to better outcomes through earlier commencement of disease-modifying treatment [23]. The findings from our study may help promote the awareness of PH as a sequalae of treatable conditions in the unique and vulnerable population of the Top End.

The burden of heart failure with preserved ejection fraction (HFpEF) is increasing [24] and it is estimated that 50 to >80% of HFpEF patients have PH [22]. Hypertension, the major cause of left ventricular diastolic dysfunction and HFpEF, was found to be very common in our study. We suspect that the prevalence of PH-LHD was underestimated due to our strict criteria in defining PH-LHD. Left atrial dilatation, an indicator of left heart pathology, was present in 48% of PH patients without an identifiable cause, reinforcing our suspicion.

The population studied exemplifies many of the difficulties of managing a rural, remote population in an era of increasing medical specialization. Many patients had multiple comorbidities and chronic diseases with multiorgan involvement requiring multiple specialists’ input. This could potentially lead to disjointed care resulting in suboptimal outcomes. A specialist multidisciplinary PH clinic with a collaborative approach would be an ideal care model and establishment of these in resource poor setting could be challenging but worth consideration. The fact that 17% of the cohort did not have an attributable cause for PH was an indication for under-recognition and under-investigation of PH in the region.

### 5.1. Future directions

Based on findings of this study, it is proposed that region-specific diagnostic algorithms and referral pathways based on current clinical guidelines could assist with early diagnosis of PH. We intend to link our data with the National Echo Database Australia (NEDA)[25, 26] which will enable us to investigate the echocardiographic screening tools in triaging patients who require further invasive investigations [6, 27]. This is highly relevant in the Top End where there are many remote patients who will benefit from such readily available, non-invasive screening methods. A population screening approach may be beneficial and should be considered in regions our study identified to have disproportionately high prevalence of PH. Potential establishment of electronic database for ongoing prospective evaluation and monitoring therapy for these high-risk patients need to be considered. The delay in diagnosis could be mitigated by development and implementation of region-specific diagnostic algorithms. Prognosis could be improved by establishment of multidisciplinary model of care for early comprehensive assessment and intervention of underlying conditions (such as timely heart valve surgery) and commencement of evidence-based therapy for PAH patients.

## 6. Conclusion

Pulmonary hypertension in the Top End of Australia was considerably more common compared to previously published community cohort studies. Indigenous patients developed PH at younger age but suffered higher mortality. Modifiable risk factors for PH were also prevalent prompting an urgent need to adopt a preventive strategy. Underutilization of advanced therapy in PAH patients as well as apparent under-investigation need further investigation and mitigation to improve outcomes.

## Data Availability

Happy to share de-indentified data if interested

